# Patterns of Patient Engagement in Antiretroviral Therapy Care: a retrospective cohort study in Malawi

**DOI:** 10.1101/2025.04.22.25326196

**Authors:** Hannock Tweya, Agness Thawani, Jacqueline Huwa, Ethel Rambiki, Evelyn Viola, Layout Gabriel, Geldert Chiwaya, Joseph Chintedza, Aubrey Kudzala, Christine Kiruthu-Kamamia, Pachawo Bisani, Marrianne M Holec, Caryl Feldacker

**Affiliations:** Department of Global Health, University of Washington, Seattle, WA, USA; International Training and Education Center for Health (I-TECH), Seattle, WA, USA; Lighthouse Trust, Lilongwe, Malawi

**Keywords:** Patient engagement, Antiretroviral Therapy, Retention

## Abstract

**Background:** Consistent engagement in antiretroviral therapy (ART) care is crucial for health outcomes and HIV transmission reduction. This study examined the first two years of ART engagement patterns in two public ART clinics in Lilongwe, Malawi. Routine retention support is provided using ART “Buddies” or a two-way texting (2wT) system for those with phones and interest.

**Methods:** ART engagement patterns were analysed across six-month intervals (>0–6, >6–12, >12–18, >18–24). Patients retained on ART were categorised as *continuously engaged* (attended all appointments within 13 days), *cyclical engagement* (returned late (14–59 days) at least once), and *re-engaged* (missed an appointment by ≥60 days but returned to care). Clients who disengaged (lost to follow-up (LTFU), transferred out, stopped, or died) at any interval were assigned that outcome. Engagement patterns were visualised using a Sankey chart.

**Results:** Among 6,303 clients, 1,030 (16%) were in the Buddy support group with phone access, 4,850 (77%) without phone, and 423 (7%) in the 2wT support group. 5,300 (84%) clients were grouped into 33 common engagement patterns over 24 months; 1,003 (16%) of clients illustrated fewer common patterns. By 24 months, 3,368 (53%) were retained on ART: 296 (70%) of 2wT; 2,793 (58%) of Buddy with phone access; and 3,367 (27%) of Buddy without phone access. Among the 3,368 clients retained on ART at 24 months, 1,836 (55%) were continuously engaged, 1,031 (30%) had cyclical engagement, and 500 (15%) re-engaged after LTFU. Overall, 1836 (29% of the total cohort of 6,303) continuously engaged in care over 24 months. In the six-month interval analysis, clients aged 50+ had the highest proportion (n=184, 70%) of continuous engagement overall, compared to individuals aged 35-49 years (n=832, 58%) and 18-34 years (n=820, 49%). 2wT clients showed the highest continuously engagement up to 18 months (70% at 0–6 months, 72% at >6–12 months, and 81% at >12– 18 months), compared to Buddy groups (with phone access:57%, 58%, 76%; without phone access: 30%, 33%, 62%).

**Conclusion:** A small proportion of clients were continuously on ART over the first 24 months of ART. Older clients and 2wT participants had more favourable ART engagement patterns; those without phone access faired worst. While most ART clients followed similar ART engagement patterns, a few showed varying trajectories. Tailored retention support based on engagement patterns could improve long-term retention in ART care.

## Introduction

Retention in antiretroviral therapy (ART) care is essential for ensuring sustained viral suppression, improving health outcomes, and preventing the onward transmission of HIV[1–3]. However, ART retention is challenging, particularly in low- and middle-income countries [4–7]. People living with HIV (PLHIV) often face well-documented structural and individual barriers to HIV care, such as economic constraints, transportation difficulties, stigma, and mental health issues [8–10]

Traditionally, ART retention, defined as actively receiving ART, has been assessed cross-sectionally where PLHIV are categorised as either ‘retained’ or ‘not retained’ at specific time points, typically at 6, 12 or 24 months after ART initiation[11–13]. This time-specific measure of ART retention has significantly shaped global policies but fails to capture the complexities of client behaviour between these time markers. With the scale-up of electronic medical records systems (EMRS) which provide comprehensive, longitudinal data on PLHIV receiving ART, there is an opportunity to map more detailed client retention in care pathways encompassing ART engagement [11].

ART engagement is dynamic[14]. PLHIV frequently cycle in and out of care as they navigate changing personal, social, and economic circumstances[13]. Previous studies illustrate that early disengagement, (lack of retention within the first six months on ART,) is particularly predictive of longer-term disengagement and loss to follow-up (LTFU)[11,15], further supporting the notion that ART engagement is not a uniform process but differs across stages and client groups[14,16].

In 2019, the Lighthouse Trust (LT), a local non-government organisation providing HIV services in Malawi, introduced a “Buddy” support service as a standard of care (SoC) at its ART clinics, Lighthouse (LH) and Martin Preuss Centre (MPC) in Lilongwe, Malawi. Buddies are PLHIV who offer peer social support, encouragement, and reminders for upcoming and missed visits during the first 12 months of ART. In 2021, LT implemented two-way texting (2wT), a hybrid intervention combining an automated weekly blast of a non-HIV-related motivational message and specific, interactive, response-requested, ART visit reminders[17]. Preliminary evidence demonstrated 2wT is effective, with a 91% ART retention rate at 12-months compared to 76% with SoC. LT also implements a SoC Back-To-Care (B2C) program, reactive late ART retention support, tracing all clients who miss appointments by 14 days or more[18]. Understanding the ART engagement patterns—given that PLHIV frequently cycle in and out of care—is crucial for refining interventions and developing additional strategies to enhance and sustain long-term retention.

The objectives of this study were: 1) Describe characteristics of ART clients and ART program outcomes by type of retention support: a) SoC Buddy among clients with phone access; b) SoC Buddy among clients without phone access; and c) 2wT; 2) Visualise ART engagement patterns over 24 months to understand how ART clients cycle in and out of care; 3) Assess ART engagement patterns by sex, age group, and type of retention support at six-month intervals following ART initiation; and 4) Describe on-time clinic visit attendance over the first 24 months on ART.

## Methods

### Study design

This retrospective descriptive cohort study utilised routine program data from the LT clinics (LH and MPC) in Lilongwe, Malawi. The study included all ART clients who initiated ART between 01 January 2020 and 31 December 2021, allowing a 24-month ART follow-up period by 31 December 2023.

### Settings

LT clinics use point-of-care (PoC) EMRS for client management[19]. Clients diagnosed with HIV are registered in the PoC EMRS and referred to an ART Buddy for pre-ART initiation counselling, tracing and psychosocial support. Clients are then assessed for their WHO HIV clinical stage. ART appointments are typically scheduled monthly for the first three months, then aligned with the six-month viral load (VL) monitoring. After six months, appointments are scheduled every three or six months, depending on client’s condition. In addition to their standard ART supply, clients receive an additional two days’ worth of ART for each appointment to ensure they do not run out of medication. Clients who transfer to LT from other ART facilities are registered in the EMRS and receive the same ART and adherence support.

### ART retention support

After pre-ART counseling, clients can choose early retention support through either SoC Buddies or the 2wT platform, described in detail previously [17]. In brief, a Buddy calls clients to remind them of their clinic appointments (if a client has a phone) and offers in-person adherence counseling during clinic visits in the first 12 months of ART. Clients who opt for 2wT must own a phone and receive a confirmatory 2wT registration message before accessing retention support. Once registered, the 2wT platform sends personalised visit reminders on days 3 and 1 before an appointment and on days 2, 5 or 11 after a missed appointment, if needed. Clients respond to visit reminders with “yes” or “no” to confirm attendance. Additionally, the platform sends weekly motivational messages on various health topics. Clients can interact via SMS with a 2wT officer during routine clinic hours. 2wT retention support was offered for up to 24 months post-ART initiation. 2wT clients do not receive Buddy support after initiation. For all clients who miss appointments for 14 days or more, B2C conducts phone calls or home visits to encourage them to return for care.

### Definitions

Clients were divided into 3 groups: Buddy support with a recorded phone number (Buddy support with phone); Buddy support without a recorded phone number (Buddy support without phone); and 2wT (phone required).

For this analysis, we defined four outcome measures: Malawi Ministry of Health (MoH) ART program outcomes, retention, ART engagement patterns, and on-time visit attendance.

MoH ART outcomes: At each visit, clients who continued treatment were classified as 1) Retained on ART; 2) LTFU: clients who missed clinic appointments for at least 60 days; 3) Stopped ART treatment: clients who discontinued treatment on their own or due to ART provider’s decisions; 4) Transfer-out: clients who moved their care to another ART facility; and 5) Died: Any cause of death.

Retention was defined as the proportion of clients retained on ART at 6-, 12-, 18- and 24-months post-initiation divided by the number of clients who started ART between 01 January 2020 and 31 December 2021.

<u>ART engagement patterns</u> aim to deepen understanding of client behaviours of cycles in and out of care and defined based on the (i) MoH ART program outcome at the end of each six-month interval (>0-6, >6-12, >12-18, and >18-24) and (ii) within-interval appointment attendance patterns. Clients who disengaged from ART care (LTFU, transfer out, stopped, or died) at the end of any interval were assigned that outcome. Clients whose outcome was retained on ART at the end of any interval were further categorised by within-interval appointment attendance pattern: (i) Continuously engaged: clients with all appointments attended within 13 days of the scheduled date; (ii) Cyclical engagement: clients who returned late (between 14 and 59 days) at least once; (iii) Re-engaged: clients who missed an appointment at least once by 60 days or more and returned to care. The least desirable engagement pattern was assigned to client retained on ART if they had multiple engagement patterns during any 6-month interval. For example, a client retained on ART at 12 months who attended two appointments on time but missed a visit by 58 days between months 6 and 12 would be assigned “cyclical engagement” at 12 months.

On-time visit attendance was categorized ‘Yes’ if a client visited within 13 days of the appointment, ‘No’ if they visited after 13 days, or they did not return at all.

### Statistical analysis

PoC EMRS data from LH and MPC were extracted between 10 March and 20 May 2024. Data were analysed using Stata version 18. Descriptive statistics (frequencies and medians with interquartile range (IQR)) were used to describe the study population. ART retention and engagement pattern proportions are presented in each six-month interval, stratified by sex, age group, and type of retention support: Buddy support with or without mobile phone and 2wT. Access to the phone was defined based on the recorded phone in the EMRs. A Z-test for proportions was used to determine which variable levels or pairwise comparisons contributed to statistical significance. Due to the large sample size, a 95% confidence interval was used to assess the significance level and p-value from the z-test was presented in the text. The chi-square test for trends was employed to assess trends in each level of the six-month outcomes (at 6, 12, 18, and 24 months) across age groups. To account for multiple levels of the six-month outcome, binary variables were created for each level.

Across the time points 6, 12, 18 and 24 months, ART engagement patterns were analysed and quantified, identifying 322 unique patterns, most of which had a single client. Overall engagement patterns were visualised using a Sankey chart, illustrating trajectories for those retained on ART (continuously engaged, cyclical engagement, re-engaged) and those who disengaged (LTFU, stopped ART, transferred-out, died) at 6, 12, 18, and 24 months. To enhance visualisations, two Sankey charts were created based on a qualitative segmentation approach: one for common patterns illustrated by ≥21 clients and another for client patterns with 20 or fewer clients.

### Ethics considerations

Ethics approval was obtained from the Malawi National Health Sciences Research Committee (Protocol #23/10/4258). Routine program data without client identifiers was used therefore participant informed consent specifically to this study was not required.

## Results

### Study population

Of 6,990 PLHIV who initiated ART between 1 January 2020 and 31 December 2021, a total of 687 (10%) were excluded because they had received ART for six months or more at other sites before enrolling at the study facilities. The remaining 6,303 (90%) were included: 1,030 (16%) in the Buddy support group with phone, 4,850 (77%) in the Buddy support group without phone, and 423 (7%) in the 2wT support group (phone required) **(Table 1)**. The type of retention support was associated with sex, age group at ART initiation, on-time visit attendance and MoH ART outcomes at 24 months (χ^2^ test, p<0.001). There were more women (58%) than men (42%), with a more notable difference among those without phone (female: 66% vs male: 34%, z-test: p< 0.001). The SoC without phone had a significantly higher proportion of clients aged 18-34 (62%) compared to those in SoC with phone (53%, z-test, p<0.001) and the 2wT group (53%, z-test, p=0.002).

**Table 1.** Baseline characteristics and outcomes of PLHIV who received ARVs between 2020 and 2023 at Lighthouse and Martin Preuss Centre, Lilongwe, Malawi.

|  | Total | Participants Groups |  |  | Chi-square test* |
| --- | --- | --- | --- | --- | --- |
|  |  | Buddy support without phone | Buddy support with phone | Two-way Texting |  |
|  | 6,303 | 1,030 (16%) | 4,850 (77%) | 423 (7%) |  |
| Sex |  |  |  |  | <0.001 |
| Male | 2,663 (42%) | 349 (34%) | 2,115 (44%) | 199 (47%) |  |
| Female | 3,640 (58%) | 681 (66%) | 2,735 (56%) | 224 (53%) |  |
| Age at ART initiation (years) |  |  |  |  | <0.001 |
| 18-34 | 3,437 (55%) | 639 (62%) | 2,574 (53%) | 224 (53%) |  |
| 35-49 | 2,419 (38%) | 325 (32%) | 1,922 (40%) | 172 (41%) |  |
| 50+ | 447 (7%) | 66 (6%) | 354 (7%) | 27 (6%) |  |
| WHO stage |  |  |  |  | 0.206 |
| 1 or 2 | 4,812 (79%) | 797 (81%) | 3,695 (79%) | 320 (76%) |  |
| 3 | 868 (14%) | 133 (14%) | 663 (14%) | 72 (17%) |  |
| 4 | 394 (6%) | 53 (5%) | 313 (7%) | 28 (7%) |  |
| With ≥ 1+ scheduled clinic |  |  |  |  | <0.001 |
| Yes | 5,892 (93%) | 922 (90%) | 4,559 (94%) | 411 (97%) |  |
| No | 412 (7%) | 108 (10%) | 291 (6%) | 12 (3%) |  |
| Overall on time attendance<sup>β</sup> |  |  |  |  | <0.001 |
| Yes | 2,396 (41%) | 198 (21%) | 1,959 (43%) | 239 (58%) |  |
| No | 3,496 (59%) | 724 (79%) | 2,600 (57%) | 172 (42%) |  |
| MoH ART outcomes at 24 months |  |  |  |  | <0.001 |
| Retained on ART | 3,367 (53%) | 278 (27%) | 2,793 (58%) | 296 (70%) |  |
| LTFU | 1,560 (25%) | 456 (44%) | 1,047 (22%) | 57 (13%) |  |
| Stop | 290 (5%) | 59 (6%) | 219 (5%) | 12 (3%) |  |
| Transfer out | 889 (14%) | 209 (20%) | 634 (13%) | 46 (11%) |  |
| Died | 197 (3%) | 28 (3%) | 157 (3%) | 12 (3%) |  |

Notes: <sup>a</sup>Chi-square test for association; MoH= Ministry of Health, WHO = World Health Organization, PLHIV = people living with HIV, ARVs = antiretrovirals; Having a phone was defined based on the availability of a recorded phone in the EMRs; <sup>b</sup>Overall on-time attendance =Attended all visits within 13 days of the appointment.

### Overall on-time visit attendance

Of the 6,303 PLHIV, 5,892 (93%) had at least one scheduled clinic visit with a median of 9 (IQR: 7-10) visits over 24 months (**Table 1**). Among those with scheduled clinic visits, 41% attended all appointments “on-time”: more in the 2wT support group (58%) compared to 43% (z-test, p<0.001) of Buddy support with phone and 21% (z-test, p<0.001) of Buddy support without phones.

### 24-months ART outcomes

At 24 months post-ART initiation, 3,367 (53%) were retained on ART and 2,936 (47%) disengaged from care: 1,560 (25%) were LTFU, 290 (5%) stopped treatment, 889 (14%) transferred out and 197 (3%) were dead (**Table 1**). The 2wT group had a significantly higher proportion of clients retained on ART (70%), followed by the Buddy support group with phone (58%, z-test: p<0.001) and those without phone (27%, z-test: p<0.001).

### ART Engagement Patterns: Overall and by Six-Month Intervals

**Figure 1a** shows ART engagement patterns for 5,300 (84%) clients, each containing at least 21 clients, over six-month intervals in the first 24 months of ART. 33 unique engagement patterns were identified, showing cycling in and out of ART care. In the first six months, 2,968 (56%) remained continuously engaged, 599 (11%) had cyclical engagement, and 870 (16%) were LTFU. Between 6 and 12 months, 2,410 (65%) were continuously engaged, with the proportion increasing to 83% between 18 and 24 months. Notably, 75 (1%) clients who were LTFU within the first six months re-engaged in care before the end of this period. Some clients with cyclical engagement at 12 months transitioned to continuous engagement at 18 or 24 months. By 24 months, 83% of clients were continuously engaged.

**Fig 1a.**
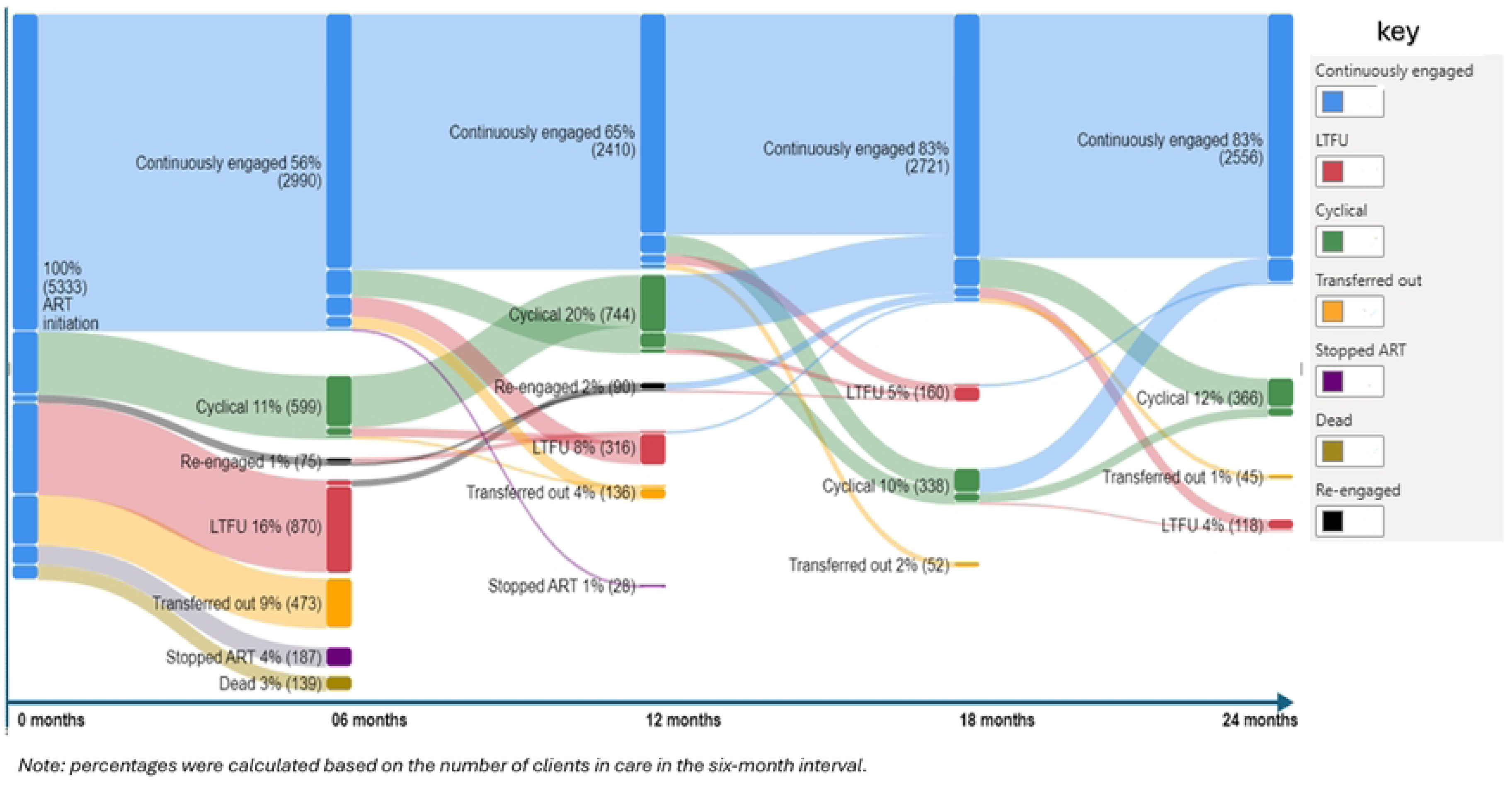
Transitions in ART engagement patterns with more than 21 clients over 24 months post ART Initiation.

**Fig 1b** shows engagement patterns involving 20 or fewer clients who were not included in **Fig 1a**. A total of 1,003 (16%) had 289 unique ART engagement patterns between months 0 and 24, exceeding the patterns shown in **Fig 1a**.

**Fig 1b.**
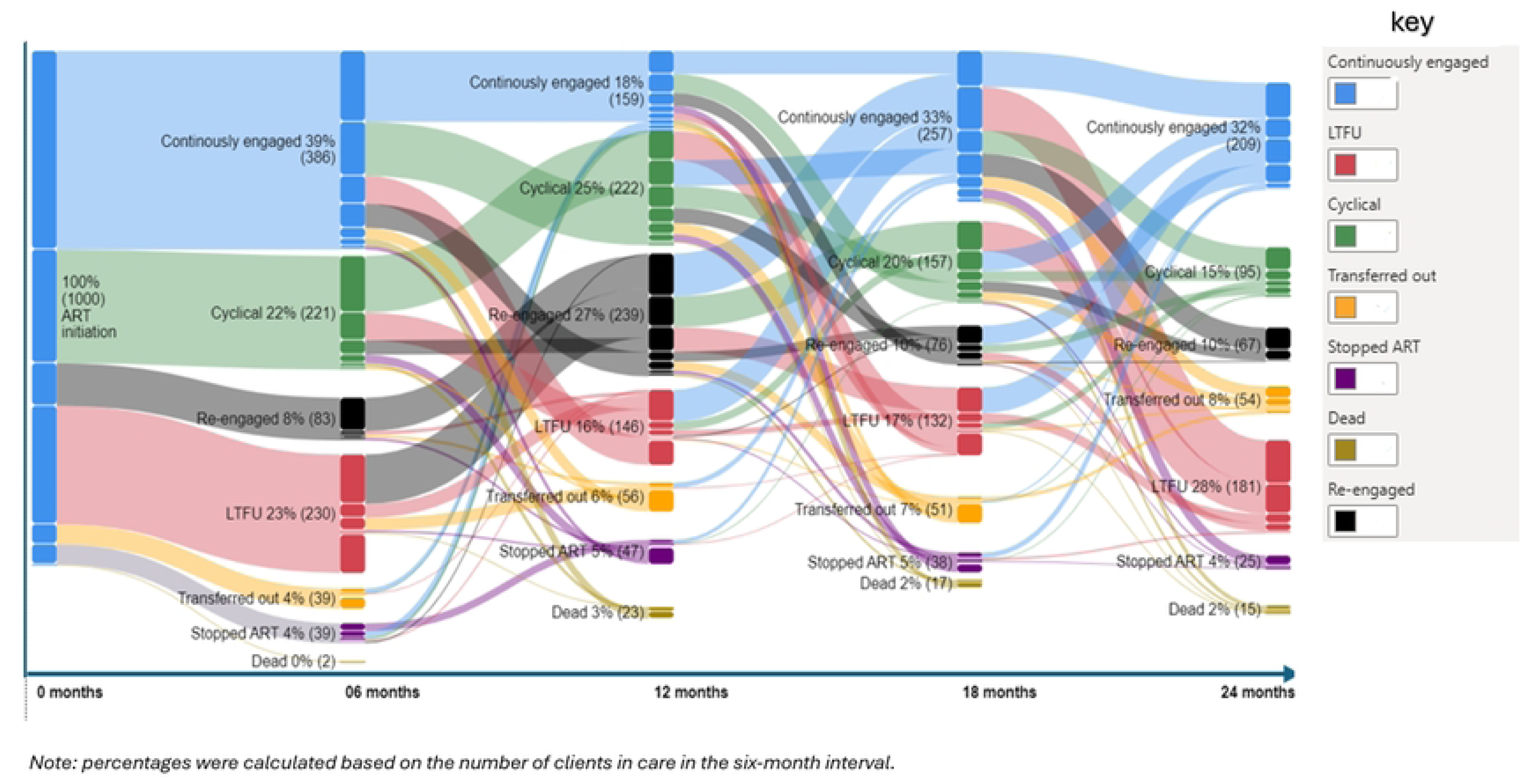
Transitions in ART engagement patterns with less than 21 clients over 24 months post ART Initiation.

### ART engagement patterns by sex

At 24 months, 1,461 (55%) males were retained on ART, compared to 1,906 (52%) females (z-test, p<0.001). (**Table 2**). In the six-month interval analysis, males were slightly more continuously engaged than females in the first six months of ART, with 80% vs 76% (z-test, p<0.001). However, after 6 months of ART, the level of continuous engagement was similar. The proportions of LTFU, cyclical engagement, and re-engagement after LTFU were similar by sex.

**Table 2.** ART engagement patterns by sex: overall and at six-month intervals (2020 and 2023) at Lighthouse and Martin Preuss Centre, Lilongwe, Malawi.

| Retention patterns | Total | Sex* |  |
| --- | --- | --- | --- |
|  |  | Male | Female |
| Total patients | 6,303 | 2,663(42%) | 3,640 (58%) |
| **Overall: 0 to 24 months** |  |  |  |
| Retained on ART | 3367 (53%) | 1461 (55%) | 1906 (52%)* |
| Continuously engaged | 1,836(55%) | 818 (56%) | 1,018(53%) |
| Cyclical | 1,031 (31%) | 436 (30%) | 595 (31%) |
| Re-engaged | 500 (15%) | 207 (14%) | 293 (15%) |
| LTFU | 1,560 (25%) | 641 (24%) | 919 (25%) |
| Stopped ART | 290 (5%) | 128 (5%) | 162 (4%) |
| Transferred out | 889 (14%) | 325 (12%) | 564 (15%) * |
| Dead | 197 (3%) | 108 (4%) | 89 (2%) |
| **0 to 6 months** |  |  |  |
| Total patients | 6,300 | 2,657 | 3,636 |
| Retained on ART | 4,321 (69%) | 1831 (69%) | 2483(68%) |
| Continuously engaged | 3,361 (78%) | 1,459 (80%) | 1,896 (76%)* |
| Cyclical | 802 (18%) | 323 (17%) | 478 (19%) |
| Re-engaged | 158 (4%) | 49 (3%) | 109 (5%) |
| LTFU | 1,100 (17%) | 457 (17%) | 643 (18%) |
| Stopped ART | 226 (4%) | 104 (4%) | 122 (3%) |
| Transferred out | 512 (8%) | 187 (7%) | 325 (9%)* |
| Dead | 141 (2%) | 78 (3%) | 63 (2%) |
| **>6 to 12 months** |  |  |  |
| Total patients | 4,614 | 1,967 | 2,647 |
| Retained on ART | 3,832 (83%) | 1618 (82%) | 2214(84%) |
| Continuously engaged | 2,575 (67%) | 1,122 (69%) | 1,453 (66%) |
| Cyclical | 928 (24%) | 364 (22%) | 564 (25%)* |
| Re-engaged | 329 (9%) | 132 (8%) | 197 (9%) |
| LTFU | 462 (10%) | 213 (11%) | 249 (9%) |
| Stopped ART | 75 (2%) | 45 (2%) | 30 (1%)* |
| Transferred out | 220 (5%) | 76 (4%) | 144 (5%) |
| Dead | 25 (1%) | 15 (1%) | 10 (<1%) |
| **>12 to 18 months** |  |  |  |
| Total patients | 4,051 | 1,721 | 2,330 |
| Retained on ART | 3,583(88%) | 1,540(89%) | 2,043(88%) |
| Continuously engaged | 3,024 (84%) | 1,285 (83%) | 1,739 (85%) |
| Cyclical | 483 (13%) | 223 (14%) | 260 (13%) |
| Re-engaged | 76 (2%) | 32 (2%) | 44 (2%) |
| LTFU | 282 (7%) | 116 (7%) | 166 (7%) |
| Stopped ART | 40 (1%) | 15 (1%) | 25 (1%) |
| Transferred out | 124 (3%) | 43 (2%) | 81 (3%) |
| Dead | 22 (0%) | 7 (0%) | 15 (1%) |
| **>18 to 24 months** |  |  |  |
| Total patients | 3,795 | 1,639 | 2,156 |
| Retained on ART | 3363(89%) | 1458(89%) | 1905(88%) |
| Continuously engaged | 2,822 (84%) | 1,230 (84%) | 1,592 (84%) |
| Cyclical | 473 (14%) | 201 (14%) | 272 (14%) |
| Re-engaged | 68 (2%) | 27 (2%) | 41 (2%) |
| LTFU | 291 (8%) | 121 (7%) | 170 (8%) |
| Stopped ART | 25 (1%) | 9 (1%) | 16 (1%) |
| Transferred out | 99 (3%) | 41 (3%) | 58 (3%) |
| Dead | 17 (0%) | 10 (1%) | 7 (0%) |

\*Statistical analysis: Pairwise comparisons were conducted using 95% confidence intervals from Z-tests with, (\*) indicates statistically significant results.

### ART engagement patterns by age group

At every six-month interval, age was associated with ART engagement (**Table 3**). Among clients retained on ART, clients ages 50 years and above had the highest proportion of continuously engaged clients (70%), with continuous engagement decreasing among clients aged 35-49 (58%) and 18-34 (49%) (χ^2^ test for trend, p <0.001). Among all study clients, LTFU was the highest in the 18-34 age group (28%) compared to 21% in the 35-49 group and 19% in the 50 and above group (χ^2^ for trend, p <0.001). Mortality was highest in the 50+ years group (9%) (χ^2^ for trend, p <0.001).

**Table 3.** ART engagement patterns by age group: overall and at six-month intervals (2020 and 2023) at Lighthouse and Martin Preuss Centre, Lilongwe, Malawi.

| Retention patterns | Age in years at ART initiation |  |  | Chi-square test for trend |
| --- | --- | --- | --- | --- |
|  | 18-34 | 35-49 | 50 and above |  |
| Total patients | 3,437 (55%) | 2,419 (38%) | 447 (7%) |  |
| **Overall: 0 to 24 months** |  |  |  |  |
| Retained on ART | 1674(49%) | 1431(59%)** | 262(59%) | <0.001 |
| Continuously engaged | 820 (49%) | 832 (58%) | 184 (70%) | <0.001 |
| Cyclical | 526 (31%) | 441 (31%) | 64 (24%) | 0.210 |
| Re-engaged | 328 (20%) | 158 (11%) | 14 (5%) | <0.001 |
| LTFU | 977 (28%) | 497 (21%) | 86 (19%) | <0.001 |
| Stopped ART | 198 (6%) | 85 (4%) | 7 (2%) | <0.001 |
| Transferred out | 524 (15%) | 312 (13%) | 53 (12%) | <0.013 |
| Dead | 64 (2%) | 94 (4%) | 39 (9%) | <0.001 |
| **0 to 6 months** |  |  |  |  |
| Total patients | 3,436 | 2,417 | 447 |  |
| Retained on ART | 2,275 (66%) | 1,741 (72%) | 305 (68%) | 0.133 |
| Continuously engaged | 1,686 (74%) | 1,410 (81%) | 265 (87%) | <0.001 |
| Cyclical | 488 (22%) | 283 (16%) | 31 (10%) | <0.001 |
| Re-engaged | 101 (4%) | 48 (3%) | 9 (3%) | <0.032 |
| LTFU | 684 (20%) | 349 (14%) | 67 (15%) | <0.001 |
| Stopped ART | 152 (4%) | 67 (3%) | 7 (2%) | <0.001 |
| Transferred out | 282 (8%) | 192 (8%) | 38 (9%) | <0.941 |
| Dead | 43 (1%) | 68 (3%) | 30 (7%) | <0.001 |
| **>6 to 12 months** |  |  |  |  |
| **Total patient** |  |  |  |  |
| Total patients | 2,475 | 1,828 | 311 |  |
| Retained on ART | 1,972(80%) | 1,577(86%) | 283 (91%) | 0.038 |
| Continuously engaged | 1,229 (62%) | 1,123 (71%) | 223 (79%) | <0.001 |
| Cyclical | 514 (26%) | 359 (23%) | 55 (19%) | <0.249 |
| Re-engaged | 229 (12%) | 95 (6%) | 5 (2%) | <0.001 |
| LTFU | 301 (12%) | 144 (8%) | 17 (5%) | <0.001 |
| Stopped ART | 53 (2%) | 22 (1%) | 0 (0%) | 0.001 |
| Transferred out | 140 (6%) | 73 (4%) | 7 (2%) | 0.015 |
| Dead | 9 (<1%) | 12 (1%) | 4 (1%) | 0.031 |
| **>12 to 18 months** |  |  |  |  |
| Total patients | 2,117 | 1,640 | 294 |  |
| Retained on ART | 1810 (85%) | 1498(91%) | 275(94%) | 0.134 |
| Continuously engaged | 1,499 (83%) | 1,270 (85%) | 255 (93%) | 0.002 |
| Cyclical | 264 (15%) | 201 (13%) | 18 (6%) | 0.047 |
| Re-engaged | 47 (3%) | 27 (2%) | 2 (1%) | 0.050 |
| LTFU | 189 (9%) | 84 (5%) | 9 (3%) | <0.001 |
| Stopped ART | 27 (1%) | 13 (1%) | 0 (0%) | 0.024 |
| Transferred out | 84 (4%) | 37 (2%) | 3 (1%) | <0.001 |
| Dead | 7 (0%) | 8 (0%) | 7 (2%) | <0.001 |
| **>18 to 24 months** |  |  |  |  |
| Total patients | 1,941 | 1,577 | 277 |  |
| Retained on ART | 1673(86%) | 1429 (91%) | 261 (94%) | 0.211 |
| Continuously engaged | 1,359 (81%) | 1,222 (86%) | 241 (92%) | 0.007 |
| Cyclical | 264 (16%) | 191 (13%) | 18 (7%) | 0.007 |
| Re-engaged | 50 (3%) | 16 (1%) | 2 (1%) | 0.608 |
| LTFU | 178 (9%) | 100 (6%) | 13 (5%) | <0.001 |
| Stopped ART | 17 (1%) | 8 (<1%) | 0 (0%) | 0.054 |
| Transferred out | 65 (3%) | 31 (2%) | 3 (1%) | 0.003 |
| Dead | 8 (0%) | 9 (0%) | 0 (0%) | 0.833 |

In the six-month interval analysis, clients aged 50 years and above had the highest proportion of continuous engagement among clients retained on ART, with 87% at 0-6 months, 79% at >6-12 months, 93% at >12-18 months, and 92% at >18-24 months, compared to 81%,71%, 85%, 86% in ages 35-49 years and 74%, 62%, 83% and 81% in ages 18-34 years, respectively (χ^2^ for trend, all p-values <0.01). Cyclical engagement was lowest in ages 50 years and above, ranging between 6% and 7%, while in other younger age groups, it was consistently above 12%.

### ART engagement patterns by type of retention support

The type of retention support was associated with ART engagement patterns in all six-month intervals (**Table 4**). Overall, the 2wT group had the highest number of retained clients (70%), followed by Buddy support with phone (58%), then Buddy support without phone (27%)(z-test, p<0.001). Sixty-eight percent of clients in the 2wT group were continuously engaged compared to 55% in the Buddy group with phones and 36% in the Buddy group without phones (z-test, p<0.001).

**Table 4.** ART engagement patterns by participant group: overall and at six-month intervals (2020–2023) at Lighthouse and Martin Preuss Centre, Lilongwe, Malawi.

|  | Participants Groups |  |  | Chi-square test* |
| --- | --- | --- | --- | --- |
|  | Buddy support without phone access<sup>β</sup> | Buddy support with phone access | Two-way texting |  |
| Patients | 1,030 (16%) | 4,850 (77%) | 423 (7%) |  |
| **Overall: 0 to 24 months** |  |  |  |  |
| Retained on ART | 278 (27%) | 2793 (58%)** | 296(70%)** |  |
| Continuously engaged | 101 (36%) | 1,533 (55%)** | 202 (68%)** | <0.001 |
| Cyclical | 114 (41%) | 859 (31%) | 58 (20%) |  |
| Re-engaged | 63 (23%) | 401 (14%) | 36 (12%) |  |
| LTFU | 456 (44%) | 1,046 (22%)** | 57 (13%)** |  |
| Stopped ART | 59 (6%) | 219 (5%) | 12 (3%) |  |
| Transferred out | 209 (20%) | 634 (13%)** | 46 (11%) |  |
| Dead | 28 (3%) | 157 (3%) | 12 (3%) |  |
| **0 to 6 months** |  |  |  |  |
| Total patients | 1,030 | 4,848 | 423 |  |
| Retained on ART | 478(46%) | 3490(72%)** | 353(84%)** |  |
| Continuously engaged | 314 (66%) | 2,751 (79%)** | 296 (84%)** | <0.001 |
| Cyclical | 120 (25%) | 632 (18%) | 50 (14%) |  |
| Re-engaged | 44 (9%) | 107 (3%) | 7 (2%) |  |
| LTFU | 346 (34%) | 720 (15%)** | 34 (8%)** |  |
| Stopped ART | 50 (5%) | 171 (4%) | 6 (1%)** |  |
| Transferred out | 135 (13%) | 356 (7%) | 21 (5%) |  |
| Dead | 21 (2%) | 111 (2%) | 9 (2%) |  |
| **>6 to 12 months** |  |  |  |  |
| Total patients | 542 | 3,706 | 366 |  |
| Retained on ART | 367(68%) | 3,131(84%)** | 334 (91%)** |  |
| Continuously engaged | 178 (48%) | 2,135 (68%)** | 262 (78%)** | <0.001 |
| Cyclical | 124 (34%) | 750 (24%) | 54 (16%) |  |
| Re-engaged | 65 (18%) | 246 (8%)** | 18 (6%) |  |
| LTFU | 113 (21%) | 334 (9%)** | 15 (4%)** |  |
| Stopped ART | 13 (2%) | 58 (2%) | 4 (1%) |  |
| Transferred out | 46 (8%) | 162 (4%)** | 12 (3%) |  |
| Dead | 3 (1%) | 21 (1%) | 1 (<1%) |  |
| **>12 to 18 months** |  |  |  |  |
| Total patients | 398 | 3,307 | 346 |  |
| Retained on ART | 317(80%) | 2953(89%)** | 313(90%) |  |
| Continuously engaged | 247 (78%) | 2,497 (85%)** | 280 (90%)* | <0.001 |
| Cyclical | 63 (20%) | 394 (13%)* | 26 (8%) |  |
| Re-engaged | 7 (2%) | 62 (2%) | 7 (2%) |  |
| LTFU | 53 (13%) | 207 (6%)** | 22 (6%) |  |
| Stopped ART | 4 (1%) | 33 (1%) | 3 (1%) |  |
| Transferred out | 20 (5%) | 97 (3%) | 7 (2%) |  |
| Dead | 4 (1%) | 17 (<1%) | 1 (<1%) |  |
| **>18 to 24 months** |  |  |  |  |
| Retained on ART | 277(80%) | 2790(89%)** | 296 (91%) |  |
| Continuously engaged | 227 (82%) | 2,341 (84%)** | 254 (86%) | 0.001 |
| Cyclical | 44 (16%) | 393 (14%) | 36 (12%) |  |
| Re-engaged | 6 (2%) | 56 (2%) | 6 (2%) |  |
| LTFU | 46 (13%) | 229 (7%)** | 16 (5%) |  |
| Stopped ART | 4 (1%) | 20 (1%) | 1 (0%) |  |
| Transferred out | 16 (5%) | 72 (2%) | 11 (3%) |  |
| Dead | 2 (0%) | 14 (0%) | 1 (0%) |  |

<sup>β</sup>Buddy support with phone access: Defined as having a phone number recorded in the Electronic Medical Records (EMR) system. \*Statistical analysis: $\chi^2$ tests were used to examine associations between types of retention support; Pairwise comparisons were conducted using 95% confidence intervals from Z-tests with, (\*) indicates statistically significant results. Comparison groups: Buddy support without phone access vs. buddy support with phone access; Buddy support with phone access vs. two-way texting support

In the six-month interval analysis, the 2wT group had the highest proportion of clients who continuously engaged up to 18 months post-ART initiation (84% at 0-6 months, 78% at >6-12 months, and 90% at >12-18 months), compared to the Buddy group with phone (79%, 68%,85%) and without phone (66%, 48%,78%) (z-test, all p-values <0.02). Clients in the Buddy group without phones consistently had the highest proportion of LTFU, decreasing from 34% at 0-6 months to 13% at >18-24 months, as compared to LTFU rates in those same two periods in the Buddy group with phones (15% to 6%) and 2wT group (8% to 6%) (z-test, all p-values <0.001).

### Monthly on-time visit attendance

Overall, monthly average on-time visit attendance varied between 80% and 90%, with medians of 90% (IQR: 86-93) for those 2wT group, 84% (IQR: 83-87) for Buddy support with phone and 76% (IQR: 74-79) for Buddy support without phone (**Fig 2**)

**Fig 2.**
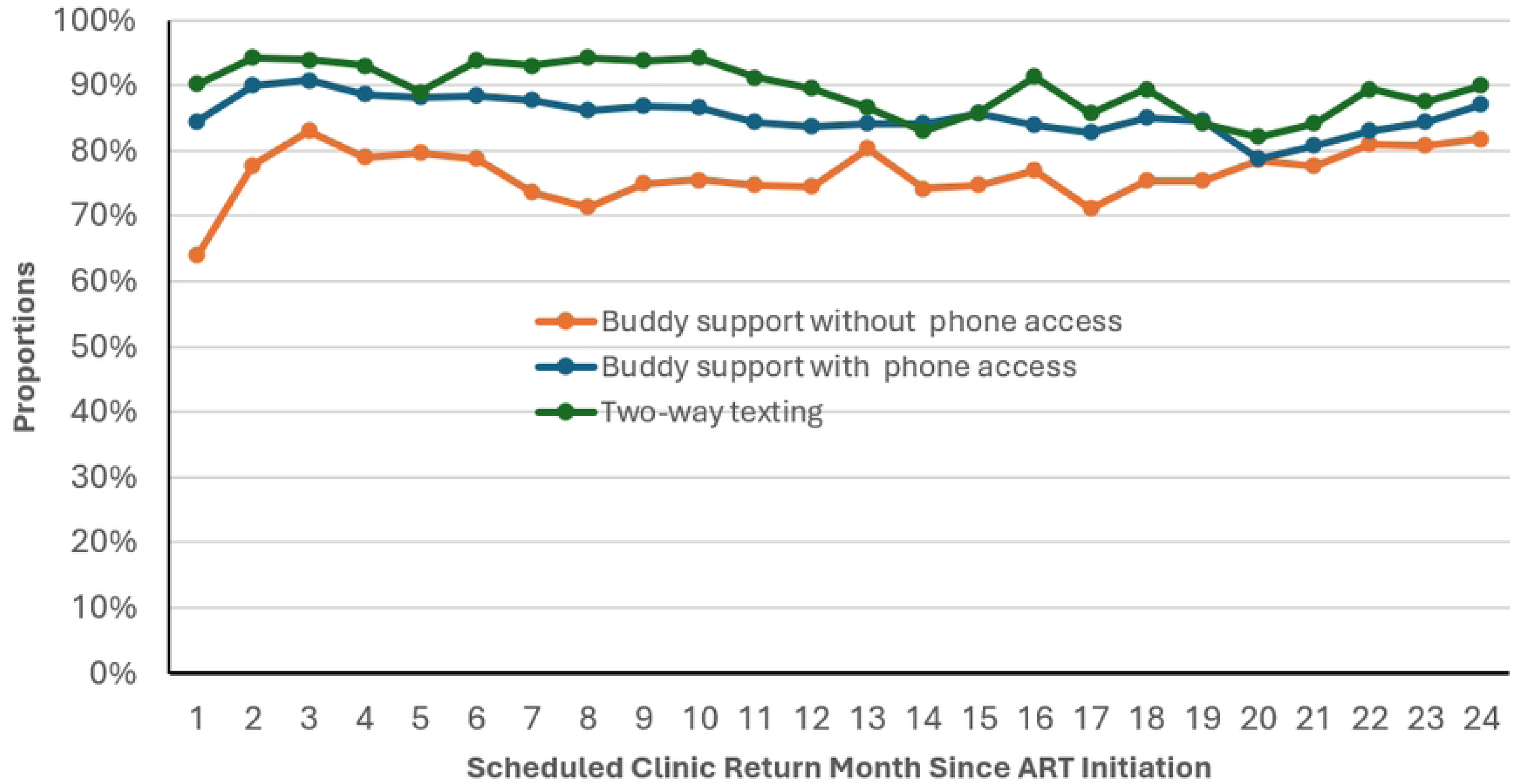
On-time visit attendance among participants expected to return to the clinic in a given month since ART initiation (2020 and 2023) at Lighthouse and Martin Preuss Centre, Lilongwe, Malawi by participants group.

## Discussion

Understanding ART engagement patterns among PLHIV is vital for viral load suppression and resource allocation. In this study, we investigated retention at six-month time-points, ART engagement patterns and on-time visit attendance trends among clients receiving ART at two large clinics in Lilongwe. Overall, 53% of clients were retained at 24 months post-ART initiation, with the highest retention observed in the 2wT group (70%) compared to those in the Buddy group with phone (58%) and no phone (27%). Of all clients, 47% disengaged (stopped ART, LTFU, transferred out and dead) from care entirely, with half of them being LTFU. Only 29% of the total cohort remained continuously engaged throughout the 24-month period, with the proportion increasing across later time points within the period. Clients aged 50+ had the highest proportion of continuous engagement. Cyclical engagement was most common among those without a phone. While most clients followed relatively consistent ART engagement, a smaller group exhibited varying engagement patterns, indicating a need for more intensive support. Only 41% attended all clinic visits on time, with the lowest attendance among those without phones (21%). We discuss several implications of these findings on practice and policy.

Cyclical engagement or re-engaged patterns are associated with poorer clinical outcomes, including risks of treatment failure and unsuppressed viral loads[20]. However, not all cyclical engagement and re-engagement instances imply treatment interruptions as some clients may obtain ART from other facilities[21], underscoring the need for improved communication with clients and transfer processes between clinics. Systems such as 2wT allow interactions between clients and retention support teams which may reduce facility tracing costs through more timely, accurate information on client ART status.

This study also highlights the complexity of clients’ ART engagement behaviours, with significant variability over the first two years of treatment. While most clients (84%) followed relatively stable trajectories with improved retention and continuous engagement over time, a smaller group (16%) had more frequent disengagement. These findings underscore the need for a differentiated approach to retention intervention, tailoring support intensity based on clients’ evolving needs. During the early phase of ART (0-6 months), universal intensive support package – including enhanced counseling, Buddy program or 2wT – can be implemented, as early retention is critical for long-term adherence. Beyond six months, once client’s engagement patterns become apparent, a stratified strategy may be appropriate. Clients with stable engagement (84%) can transition to lighter-touch interventions, such as 2wT alone, thereby serving resources. In contrast, high risk clients (16%) may require sustained and targeted support, including buddy support to prevent disengagement. Shifting from a uniform to a dynamic, data-informed retention approach can improve efficiency and optimize retention outcomes without overburdening existing systems.

Lastly, our study suggests that clients with phones had better ART engagement and on-time visit attendance than clients without recorded phone numbers. 2wT participants received two visit reminder messages before each visit and received up to three additional messages if they missed an appointment[17] while buddies contacted clients as soon as possible after missed visits. Phone access enabled earlier identification of retention problems and more timely interventions, leading to improved engagement in HIV care. The disparity in retention may also reflects deeper socioeconomic and psychosocial differences between the groups. ART clients with phones tend to have higher education levels, greater economic ability and better health literacy[22–24], all of which can improve the ability to engage with healthcare services [25]. Additionally, those who provided their phone numbers may also be more likely to have disclosed their status while those without recorded phone numbers may face more stigma or fear violence.

### Strengths and limitations

This is among the few studies to examine retention as a dynamic and evolving process to deepen understanding of how clients engage in care. Further, examination of these behaviours by both recorded phone number (a proxy for ownership or access) and proactive retention support choice is also novel in the literature, helping add knowledge that may spur retention support improvements. Our study also had limitations. First, reliance on phones can be problematic for a number of reasons. Although phone numbers confirmed at 2wT enrollment, some were deactivated or did not work consistently throughout the study. The influence of phone for SoC clients may be overstated as clients may have changed numbers over time. Clients categorized as without phones could have phones but not want to provide a number due to privacy concerns. These phone-based factors could have caused errors in client categorization, potentially influenced ART engagement, or reduced B2C success. Second, the study data were limited to two large urban sites, so cyclical and re-engagement patterns may differ elsewhere, including in smaller, rural or peri-urban clinics. Third, while utilising a large study population offers several advantages, some statistically significant differences of less than 5% between groups may not be actionable for practice or policy change.

## Conclusion

Over the first two years at these large public clinics with retention support, 28% of ART clients were continuously engaged. Older clients and clients with access to mobile phones had more favourable 24-month ART outcomes and engagement patterns over clients without documented phone numbers. Clients who opted into 2wT did better across all retention periods, suggesting that this low-cost method should be scaled as part of differentiated retention support. While most ART clients had consistent engagement patterns, with improvements in continuous engagement over time, a smaller proportion of clients showed variable ART engagement trajectories, suggesting the need for targeted retention efforts. Implementing differentiated retention strategies, with more intense support for clients based on retention risk, can improve long-term ART engagement while minimising resource waste.

## Data Availability

Data cannot be shared publicly because of include individual patient level records. Data are available from the Lighthouse Trust (contact via) for researchers who meet the criteria for access to confidential data.

## Authors’ contributions

Conceptualisation: Hannock Tweya, Caryl Feldacker

Data curation: Pachawo Bisani, Jacqueline Huwa, Agness Thawani, Ethel Rambiki, Evelyn Viola, Layout Gabriel, Geldert Chiwaya, Joseph Chintedza, Aubrey Kudzala

Formal Analysis: Hannock Tweya, Caryl Feldacker

Funding acquisition: Caryl Feldacker, Hannock Tweya

Investigation: Hannock Tweya, Caryl Feldacker, Jacqueline Huwa, Christine Kiruthu-Kamamia, Agness Thawani, Geldert Chiwaya, Joseph Chintedza, Aubrey Kudzala, Evelyn Viola, Layout Gabriel

Methodology: Hannock Tweya, Caryl Feldacker, Christine Kiruthu-Kamamia,

Project administration: Hannock Tweya, Caryl Feldacker, Ethel Rambiki, Marrianne M Holec Validation: Hannock Tweya, Caryl Feldacker, Marrianne M Holec, Pachawo Bisani Visualisation: Hannock Tweya, Caryl Feldacker

Writing-original draft: Hannock Tweya, Caryl Feldacker, Jacqueline Huwa, Agness Thawani, Layout Gabriel

Writing – Review & Editing: Pachawo Bisani, Ethel Rambiki, Evelyn Viola, Geldert Chiwaya, Joseph Chintedza, Aubrey Kudzala, Christine Kiruthu-Kamamia, Marrianne M Holec

## Acknowledgements

The authors would like to thank all the staff who collected data at Lighthouse and Martin Preuss Center clinics. We thank numerous donors supporting the Lighthouse and Martin Preuss Center clinics.

